# The impact of virtual reality cognitive behavioral therapy on mental disorders among children and youth: a systematic review and meta-analysis

**DOI:** 10.1101/2025.08.20.25334071

**Authors:** Madeline Li, Jamin Patel, Sheriff Ibrahim, Tarun Reddy Katapally

**Author notes:** Corresponding author: Tarun Reddy Katapally, (TRK).

## Abstract

**Background:** Cognitive behavioral therapy (CBT) is an effective treatment for mental disorders, however, it can be associated with limited patient engagement, low adherence, and stigma among younger populations. Virtual reality (VR) environments can facilitate innovative approaches to enhance CBT implementation in a controlled and immersive way.

**Objectives:** This study evaluates the impact of VR-CBT interventions on mental disorders in children and youth through a systematic review and meta-analysis.

**Methods:** A search was conducted in PsycINFO, PubMed, EMBASE, Scopus, and Web of Science. Studies compared VR-CBT interventions to traditional therapy or control conditions. Extracted data included post-intervention means, standard deviations, and 95% confidence intervals. Pooled effect sizes were calculated using Hedges’ g and analyzed with a random-effects model. Risk of bias was evaluated using the Cochrane risk-of-bias (RoB) 2 tool and the JBI Critical Appraisal Tool.

**Results:** In total, 20 studies were included in the systematic review, with 85% (n = 17) utilizing virtual reality exposure therapy (VRET), and 15% (n = 3) implementing broader VR-CBT frameworks. VR technologies included wearable head-mounted displays (70%, n = 14), with 30% (n = 6) relying on non-wearable systems, and 15% (n = 3) incorporating gamification elements. Seven studies were included in a meta-analysis, which showed that VR-CBT was associated with a small to moderate reduction in mental disorder symptoms in full-scale studies (pooled Hedge’s g = -0.46 (95% CI: [-0.84], [-0.09]).

**Conclusions:** VR-CBT interventions demonstrate potential for addressing mental disorders in children and youth, particularly when traditional therapy alone is insufficient and/or inaccessible.

## 1. Introduction

Mental disorders represent a major public health concern, especially among children and youth aged 24 years or younger (Sawyer et al., 2018) who exhibit notably high prevalence rates (Duncan et al., 2022; Edwards et al., 2023; Piao et al., 2022; Polanczyk et al., 2015; Zolopa et al., 2022). For instance, research shows approximately one in seven adolescents experience a mental disorder annually (WHO, 2024), significantly affecting their development (CDC, 2024b), educational outcomes (Mitchell et al., 2022), and overall well-being (CDC, 2024a). Furthermore, many mental disorders that emerge during youth persist into adulthood, with evidence indicating that around half of all lifetime mental disorders develop by the age of 14 (NAMI, n.d.). Additionally, mental disorders impose a significant global economic burden, with annual costs estimated at $1 trillion (Health, 2020). This highlights the critical need for innovative therapeutic approaches to tackle the distinct mental health challenges encountered by children and youth.

Cognitive behavioral therapy (CBT) is a well-established method for treating a range of mental disorders. CBT is grounded in cognitive theory, which posits that negative thought patterns contribute to emotional distress and behavioral problems (Chand et al., 2024; Fenn & Byrne, 2013; Zohuri & McDaniel, 2022). Through structured sessions, CBT helps individuals identify harmful thought patterns, evaluate their validity, and adopt healthier behaviors (Hofmann et al., 2012). Moreover, CBT includes various subtypes of therapeutic approaches, including exposure therapy (NHS, 2021), trauma-focused CBT (Ramirez de Arellano et al., 2014), and behavioral activation therapy (Uphoff et al., 2020), demonstrating effectiveness in treating anxiety disorders (Carpenter et al., 2018), specific phobias (Plaisted et al., 2021), depression disorders (Gautam et al., 2020), and post-traumatic stress disorder (PTSD) (Plaisted et al., 2021). Although considered a gold-standard therapeutic intervention (David et al., 2018), CBT presents multiple limitations. For instance, CBT demands active engagement and a strong commitment from individuals undergoing treatment (Institute for Quality and Efficiency in Health Care (IQWiG), 2022), which can pose challenges, particularly for children and youth. Additionally, the conventional face-to-face format may not effectively engage younger individuals, particularly when addressing certain phobias or anxieties that require controlled and practical exposure scenarios, which can be challenging to replicate during therapy sessions (Lange et al., 2012; Wallach et al., 2009). Furthermore, pursuing traditional face-to-face therapies, such as CBT, is often associated with stigma, particularly among children and youth (Sheikhan et al., 2023), who may hesitate to seek mental health care due to societal perceptions and potential judgment from peers and family (Aguirre Velasco et al., 2020; Gulliver et al., 2010; Radez et al., 2021). These challenges underscore the importance of developing more flexible and engaging therapeutic strategies to effectively support children and youth with mental disorders.

Virtual reality cognitive behavioral therapy (VR-CBT) offers a promising approach to overcoming these limitations. This cutting-edge intervention leverages advanced immersive technologies, to offer individuals a controlled and secure virtual environment for addressing and managing their mental disorders (Pons et al., 2022). VR-CBT interventions expand upon foundational CBT principles by utilizing the immersive capabilities of VR to simulate realistic scenarios. These environments enable individuals to practice coping strategies, gradually reduce sensitivity to anxiety-inducing stimuli, and benefit from real-time feedback provided by therapists (Gonçalves et al., 2012; Jeppesen et al., 2022; Riva & Serino, 2020; Wiley et al., 2022). The immersive capabilities of VR-CBT facilitate the integration of thoughts, behaviors, and emotions by immersing individuals in realistic scenarios where they can actively apply learned skills, thereby enhancing the effectiveness of therapy (Loenen et al., 2022; Wu et al., 2021).

Moreover, prior research indicates that VR-CBT interventions are more well-received and result in greater satisfaction compared to traditional therapy approaches (Beck et al., 2007; Garcia-Palacios et al., 2007). Among individuals with specific phobias, the refusal rate for VRET was only 3%, markedly lower than the 27% observed with in-vivo exposure therapy (Garcia-Palacios et al., 2007). A study exploring soldiers’ perspectives on technology-based mental health treatments found that 19% of those unwilling to consult a counselor in person expressed a willingness to engage with VR-based interventions. This suggests that VR approaches could help address specific barriers to accessing mental health care (Wilson et al., 2008). By incorporating advanced technologies, VR-CBT has the potential to enrich the traditional CBT framework, offering more engaging therapeutic experiences for children and youth (Ong et al., 2024).

Advancements in VR technology have been utilized to treat a range of mental disorders in children and youth. For example, VR therapy has been applied to manage anxiety disorders, especially specific phobias, where traditional exposure therapy may be challenging to implement (Kothgassner & Felnhofer, 2021). Current evidence indicates that VR interventions are both effective and well-accepted as a treatment method for managing psychological distress in adolescents (Kelson et al., 2021). Additionally, in hospital settings, VR has been employed as an engaging therapeutic tool to reduce pain and anxiety in adolescent inpatients (Ridout et al., 2021). However, evidence regarding the benefits of VR remains inconclusive. While some studies emphasize advantages such as enhanced emotional, social, cognitive, and motivational development, others point to potential risks, including emotional distress, anxiety, addiction, obesity, cyber sickness, and sleep disturbances (Kaimara et al., 2022). Considering the dual impact of VR interventions for mental disorders, the high prevalence of these conditions among children and youth (CDC, 2024a; NAMI, n.d.; WHO, 2024), and the challenges associated with traditional CBT (Aguirre Velasco et al., 2020; Gulliver et al., 2010; Institute for Quality and Efficiency in Health Care (IQWiG), 2022; Radez et al., 2021; Sheikhan et al., 2023), it is essential to examine the effectiveness of VR-CBT within this population.

Currently, no systematic reviews have specifically investigated the impact of VR-CBT on mental disorders in children and youth. Previous reviews have evaluated the effectiveness of VR-CBT interventions, but their focus has largely been on general populations rather than this specific demographic (Dhunnoo et al., 2024; Freitas et al., 2021; Paul et al., 2024). While some reviews have examined a range of VR interventions in children and youth, their focus has been largely limited to specific mental disorders like anxiety (Kothgassner & Felnhofer, 2021) or schizophrenia (Dellazizzo et al., 2019), rather than assessing the wider scope of VR-CBT applications. Additionally, many systematic reviews have focused on other types of VR therapies, such as VR-based distraction and relaxation techniques (Eijlers et al., 2019; Nordgård & Låg, 2021; Tas et al., 2022), instead of VR-CBT. Thus, this systematic review and meta-analysis quantitatively assesses the impact of VR-CBT interventions on symptoms of mental disorders among children and youth. Furthermore, this review will offer recommendations for the evaluation, implementation, and future development of VR-CBT targeting mental disorders in children and youth.

## 2. Methods

This systematic review and meta-analysis was conducted in accordance with the Preferred Reporting Items for Systematic Reviews and Meta-Analyses (PRISMA) 2020 guidelines (Figure S1).The methodology for this systematic review and meta-analysis was conducted in accordance with a previous protocol published in PLOS ONE (Li et al., 2025). This study was registered with PROSPERO (registration number: CRD42024593054).

Amendments were made between the registered protocol and the final study. While the protocol documented a scope assessing extended reality cognitive behavioral therapy (XR-CBT, which includes virtual, augmented, and mixed reality) the final review focused exclusively on VR-CBT interventions for mental disorders. Although the protocol planned for a priori meta-regression and subgroup analyses, these were not conducted in our review due to limited number of studies included in our analyses. Due to limited RCTs, the review included feasibility and pilot studies as part of the broader category of quasi-experimental designs already permitted in the protocol, to support a more comprehensive synthesis of the evidence. Moreover, the final review employed the JBI Critical Appraisal Tool to assess risk of bias in quasi-experimental studies, a method not specified in the protocol.

### 2.1.1. Research question

To what extent do VR-CBT interventions impact the symptoms of mental disorders in children and youth compared to traditional therapies and/or a control (e.g., no treatment)?

### 2.1.2 Hypothesis

VR-CBT shows a greater effect on symptoms associated with mental disorders in children and youth compared to traditional therapies and the control condition (e.g., no treatment) at post-intervention.

### 2.1.3 Eligibility criteria

This systematic review and meta-analysis includes studies involving children and youth ages 24 years and younger (Sawyer et al., 2018). To be eligible for inclusion, a diagnosed mental disorder or exhibit symptoms of a mental disorder (e.g., anxiety, depression, attention-deficit/hyperactivity disorder (ADHD), PTSD, etc.) is necessary. This review adopts an inclusive definition of mental disorders, aligned with the criteria outlined in the Diagnostic and Statistical Manual of Mental Disorders, Fifth Edition (DSM-5), the standard classification system used by mental health professionals (Edition, 2024). The DSM-5 provides specific criteria for diagnosing mental disorders, including mood disorders (e.g., depression), anxiety disorders (e.g., generalized anxiety disorder, phobias, and social anxiety), and other conditions such as ADHD, obsessive-compulsive disorder (OCD), and PTSD (Edition, 2024). Each disorder is defined by a distinct set of symptoms and diagnostic criteria, and this review will account for the variability of these conditions when evaluating the potential applications of VR-CBT across various mental disorders. No restrictions are placed on the severity or period of mental disorder. Mental disorder outcomes are assessed using various instruments, including the Perceived Stress Scale (PSS) (Harris et al., 2023; Lee, 2012), The Patient Health Questionnaire-9 (PHQ-9) (Costantini et al., 2021; Kroenke et al., 2001), and the Generalized Anxiety Disorder scale (GAD-7) (Johnson et al., 2019; Sapra et al., 2020). This review focuses on VR-CBT interventions, which utilize immersive technologies to deliver CBT. Included studies must evaluate the effects of VR-CBT on mental disorders. The intervention is compared to alternatives such as stand-alone CBT, other traditional therapies, or control conditions, including no treatment or placebo.

This review focuses on experimental study designs, including randomized controlled trials (RCTs), quasi-randomized controlled trials (quasi-RCTs), and controlled clinical trials (CCTs), that investigate the effects of VR-CBT interventions among children and youth and their mental disorders. Our study also included both feasibility studies and full-scale studies. Feasibility studies primarily assessed the acceptability, implementation, and preliminary efficacy of the intervention. Full-scale studies focused on evaluating the efficacy or effectiveness of the intervention using more rigorous study designs. Studies that solely explore participants’ perceptions or understanding of VR-CBT, as well as those not directly assessing its impact on mental disorders (e.g., developmental studies), are excluded. Only articles published in English are included. To ensure the review reflects current technological advancements, it has examined studies published over the last decade (January 2014 to June 2024). **Table 1** outlines the inclusion and exclusion criteria in detail.

**Table 1:** Inclusion and exclusion criteria for this systematic review.

| Concept | Inclusion | Exclusion |
| --- | --- | --- |
| Age | 24 years or younger | Studies involving participants outside the specified age range of children and youth (24 years or younger) |
| Therapeutic intervention | Studies utilizing VR-CBT to treat mental disorders in children and youth, with VR technology as a core component of the intervention (e.g., VR-CBT, VRET) | Studies employing non-immersive CBT, non-CBT therapies delivered through VR, or those concentrating solely on feasibility or user experience without assessing the impact on mental disorders |
| Comparator(s) | Studies eligible for inclusion must compare VR-CBT with traditional therapies, such as in-person therapy or stand-alone CBT. Studies incorporating a control group, including those receiving no treatment, a placebo, or standard care, will also be considered. | Studies that do not compare VR-CBT interventions with an alternative therapy or a control group will be excluded. |
| Mental disorders | Change in symptoms of mental disorder | Studies focusing on physical disorders, other health issues unrelated to mental disorders, or economic, cognitive, and physical outcomes not associated with mental disorders |
| Language | English | Studies published in languages other than English |
| Publication status | Peer-reviewed literature | Grey literature |
| Date of publication | January 2014 to June 2024 | Studies published outside the specified date range of January 2014 to June 2024 |
| Type of study | Studies employing experimental designs, such as RCTs, quasi-RCTs, and CCTs, that evaluate the efficacy of VR-CBT interventions in treating mental disorders. | Observational studies, developmental studies, or any studies that do not employ experimental or quasi-experimental designs. |

### 2.2 Study Records

#### 2.2.1 Data sources

The review draws on several databases to identify studies: Embase, PubMed, Web of Science, PsycInfo, and Scopus. Embase and PubMed provided access to biomedical research, including clinical studies on mental disorders and medical devices. PsycInfo contributes extensive literature from psychology and behavioral sciences. To reflect the interdisciplinary scope of this review, incorporating psychology, medicine, and technology, Scopus and Web of Science offer a diverse range of perspectives across multiple fields. Advanced search functionalities have been employed in all databases to ensure comprehensive and accurate retrieval of relevant studies.

#### 2.2.2 Search strategy

To ensure credibility, the search strategy was developed in consultation with domain-specific librarians and incorporated three thematic categories: therapeutic intervention, mental disorder, and age. A key concepts map is presented in Table 2. To comprehensively capture relevant literature, related concepts to VR-CBT were included in the search strategy to ensure broad coverage of interventions utilizing VR in CBT. Additionally, the reference lists of included peer-reviewed studies were manually reviewed to identify any potentially relevant articles missed during the initial search. The complete search strategy for all databases is detailed in Figure S1.

**Table 2:** Key concepts map.

| Thematic category | Search terms |
| --- | --- |
| Therapeutic intervention | Virtual Reality Cognitive Behavioral Therapy, Virtual Reality Cognitive Behavioral Therapy, Virtual Reality CBT, VRCBT, Virtual Reality Exposure Therapy, VRET, Virtual reality training, cognitive behavior* therapy with virtual reality, Augmented Reality with Cognitive Behavioral Therapy, Augmented Reality with Cognitive Behavioral Therapy, AR-CBT, |
|  | ARET, Mixed Reality with Cognitive Behavioral Therapy, Mixed Reality with Cognitive Behavioral Therapy, MR-CBT, extended reality with cognitive behavioral therapy, extended reality with cognitive behavioral therapy, XR-CBT, XRET |
| Mental disorder | Mental Health, Psychological Wellbeing, Wellbeing, Anxiet*, Phobia, OCD, Depress*, PTSD, Disorder*, Behaviour*, Behavior, Psycholog* |
| Age | Youth, Adolescent*, Teen*, Student*, Child*, college student* |

#### 2.2.3 Data screening and extraction

Following the implementation of the search strategy across the databases, the results were imported into Covidence software for systematic screening and management. In the first stage, duplicate entries were eliminated, and the remaining studies were independently evaluated by two reviewers based on their titles and abstracts to determine eligibility. Discrepancies between the reviewers were resolved through discussion. During the second stage, the full texts of the studies identified in the initial screening were carefully reviewed by both reviewers. Any studies deemed ineligible were discussed prior to proceeding with data extraction. To assess the risk of bias in the included studies, the Cochrane RoB 2 tool was utilized during the review process (Higgins et al., 2011). The Cochrane RoB 2 tool was used by one reviewer to assess potential biases in areas such as randomization, allocation concealment, blinding, incomplete outcome data, and selective outcome reporting (Higgins et al., 2011). Additionally, the JBI Critical Appraisal Tool was used to assess risk of bias in quasi-experimental studies (Munn et al., 2023). Covidence was used to present the results through the PRISMA 2020 four-phase flow diagram (Rethlefsen & Page, 2022). Covidence was also used to systematically extract data from each included study, covering categories such as study design, year of publication, study aim, sample size, baseline characteristics of the study population, type of VR-CBT intervention, and the mental disorder addressed. This study did not involve the collection of primary data, eliminating the need for ethics approval.

#### 2.2.4 Data synthesis and analysis

All data analyses and statistical modeling were performed using R 4.3.3 (RStudio Team, 2020). Post-intervention means, standard deviations (SDs), and 95% confidence intervals were extracted from each study, and effect sizes were calculated using Hedges’ g, an estimator that accounts for small sample bias (Taylor & Alanazi, 2023). Hedges’ g is particularly suitable for this study due to the potential inclusion of smaller studies. Additionally, RCTs were given greater weight, as they are widely regarded as the gold standard for minimizing selection bias through randomization and maintaining controlled conditions (Higgins et al., 2019; Phillips et al., 2022). Conversely, quasi-experimental studies may be prone to biases, particularly selection bias, as they do not utilize random assignment (Shadish et al., 2002), and performance bias, arising from inconsistent delivery of interventions across non-randomized study groups. (Armijo-Olivo et al., 2022). A random-effects model was employed to pool the calculated effect sizes across studies. This approach was chosen to account for variations both between studies and among individuals, making it well-suited for managing the heterogeneity anticipated across the included studies (Dettori et al., 2022). In contrast to a fixed-effects model, which presumes a uniform effect size across all studies, the random-effects model allows for variability in effect sizes due to differences in study populations, designs, and interventions (Dettori et al., 2022).

## 3.0 Results

### 3.1 Study selection

The systematic literature search identified a total of 2469 studies through database searches. After removing 351 duplicate records, 2118 studies remained for the title and abstract screening. Following screening, 2072 articles were excluded based on relevance, leaving 44 records for the full text screening and review. Among these studies, 24 were excluded due to ineligible outcomes, interventions, study design, patient population, as well as duplications. Thus, 20 studies met the inclusion criteria for the systematic review, along with 7 of those studies meeting the inclusion criteria for the meta-analysis (Azimisefat et al., 2022; Kahlon et al., 2023).

### 3.2 Study characteristics

**Table 3** presents the data extraction summarizing key characteristics of the included studies, including mental disorder outcome, location, population, inclusion criteria, follow up time, study group characteristics, and type of intervention. Among the 20 eligible studies, all studies had either an RCT study design (35%, n = 7) (Azimisefat et al., 2022; L. Chu et al., 2023; Kahlon et al., 2023; Maskey et al., 2019; Minns et al., 2019; Nazligul et al., 2019; Porras-Garcia, Serrano-Troncoso, Carulla-Roig, Soto-Usera, Ferrer-Garcia, Fernandez-Delcastillo Olivares, et al., 2020), or quasi-experimental design (6%, n= 13) (De Luca et al., 2021, p. 2; Farrell et al., 2021; Kahlon et al., 2019; Maskey et al., 2014; Miller et al., 2023; Orena & Baruffi, 2023; Porras-Garcia, Serrano-Troncoso, Carulla-Roig, Soto-Usera, Ferrer-Garcia, Figueras-Puigderrajols, et al., 2020; Ramsey et al., 2023; Rodero & Larrea, 2022; Sülter et al., 2022a; Y. L. Tan et al., 2025a; Wechsler et al., 2024; Whiteside et al., 2020). Among RCT studies, 5 studies were full-scale (71%) (77–81), and 2 were feasibility studies (29%) (Maskey et al., 2019; Porras-Garcia, Serrano-Troncoso, Carulla-Roig, Soto-Usera, Ferrer-Garcia, Fernandez-Delcastillo Olivares, et al., 2020). Quasi-experimental studies also included 1 case study (Orena & Baruffi, 2023), 1 case report (Porras-Garcia, Serrano-Troncoso, Carulla-Roig, Soto-Usera, Ferrer-Garcia, Figueras-Puigderrajols, et al., 2020), and 1 multiple baseline case series (Farrell et al., 2021). Quasi-experimental studies were further classified into full-scale (n=2) (Maskey et al., 2014; Rodero & Larrea, 2022), feasibility studies (n=7) (De Luca et al., 2021; Orena & Baruffi, 2023; Porras-Garcia, Serrano-Troncoso, Carulla-Roig, Soto-Usera, Ferrer-Garcia, Fernandez-Delcastillo Olivares, et al., 2020; Porras-Garcia, Serrano-Troncoso, Carulla-Roig, Soto-Usera, Ferrer-Garcia, Figueras-Puigderrajols, et al., 2020; Ramsey et al., 2023; Y. L. Tan et al., 2025a; Whiteside et al., 2020), pilot studies (n=1) (Wechsler et al., 2024), proof of concept (n=1) (Sülter et al., 2022b), and a combination of pilot and feasibility studies (n=2) (Farrell et al., 2021; Kahlon et al., 2019). The number of participants within each study had a range from (n=1) to (n=100). Moreover, studies originated from various countries, such as Spain (n=3) (Porras-Garcia, Serrano-Troncoso, Carulla-Roig, Soto-Usera, Ferrer-Garcia, Fernandez-Delcastillo Olivares, et al., 2020; Porras-Garcia, Serrano-Troncoso, Carulla-Roig, Soto-Usera, Ferrer-Garcia, Figueras-Puigderrajols, et al., 2020; Rodero & Larrea, 2022), The Netherlands (n=1)(Sülter et al., 2022b), Iran (n=1) (Azimisefat et al., 2022), China (n=1) (L. Chu et al., 2023), Norway (n=2) (Kahlon et al., 2019, 2023), the USA (n=4) (De Luca et al., 2021; Miller et al., 2023; Minns et al., 2019; Whiteside et al., 2020), the United Kingdom (n=2) (Maskey et al., 2014, 2019), Turkey (n=1) (Nazligul et al., 2019), Australia (n=1) (Farrell et al., 2021), Germany (n=1) (Wechsler et al., 2024), Singapore (n=1) (Y. L. Tan et al., 2025b), and Italy (n=2) (De Luca et al., 2021; Orena & Baruffi, 2023). The age range for child and youth participants ranged from 3-24 years old. Follow-up times after the intervention ranged from immediately after administration to 12-16 months after administration.

**Table 3.** Data extraction table summarizing key characteristics of the included studies, including mental disorder outcome, location, population, inclusion criteria, follow up time, study group characteristics, and type of intervention.

| Reference | Study design | Stage of study | Outcome (measure) | Location | Population | Inclusion criteria | Follow up time | Study groups | Intervention |
| --- | --- | --- | --- | --- | --- | --- | --- | --- | --- |
| Azimisefat et al., 2022 | RCT | Full-scale study | Acrophobia (Severity measure for acrophobia, SMA) | Iran | 16-18 years | SMA score greater than or equal to 50<br>met all diagnostic criteria on the Structured Clinical Interview for DSM-5 (SCID-5)<br>Provided written informed consent | 1 week after | VRET (n=15), mean age (SD) = 17.20 (0.77), males = 0%, females = 100%<br>Eye Movement Desensitization and Reprocessing Therapy (EMDR) (n=15), mean age (SD) = 16.93 (0.88), males = 0%, females = 100%<br>Wait list control (n=15), mean age (SD) = 16.93 (0.79), males = 100%, females = 100% | VRET group: Used VR headset to confront height-related scenarios<br>utilized Oculus Rift DK2 virtual reality headset<br>EMDR group: followed an eight-phase EMDR protocol, addressing distressing memories related to height anxiety<br>Wait list control group: received no intervention |
| Chu et al., 2023 | RCT | Full-scale study | ADHD (Attention-Deficit/Hyperactivity Disorder) | China | 3-6 years | age 3-6 years<br>IQ > 70 | 20 weeks | Intervention Group (n=39), mean age (SD) = 5 (0.82), boys = 80%, girls = 21% | Intervention group: received VR-CBT (immersive interactive video games) and Learning Style Profile |
|  |  |  | Rating Scale-IV (ADHD-RS-IV) |  |  | Able to understand instructions<br>no recent engagement in other training courses. |  | Control Group (n = 39), mean age (SD) = 5.03 (0.83), boys = 77%, girls = 23% | (LSP) training (identifying each child's learning style to better support social and behavioral skills)<br>Used a Lenovo desktop computer and a 3 m × 3 m LED floor tile screen with P3.91 resolution, enclosed by guardrails and walls<br>Control group: Only received the LSP training |
| Kahlon et al., 2023 | RCT | Full-scale study | Public speaking anxiety<br>(Public Speaking Anxiety Scale (PSAS)) | Norway | 13-16 years | 13-16 years<br>Score of 55 or more on PSAS<br>Must answer yes to both of the following: afraid of public speaking and | 3 months after | VRET + No Additional Intervention (VRET + NA) (n=16), mean age (SD) = 14.40 (1.05), male = 20%, female = 80%<br>VRET + Online Exposure Program (VRET + online EXP) (n | VRET + NA: Received VRET, practiced speech exercises in front of a virtual classroom setting, utilizing the Oculus Quest 1 headset<br>VRET + online EXP: received VRET followed by online exposure program, constructed exposure training tasks using the SMART goal |
|  |  |  |  |  |  | avoidance of public speaking when possible.<br>Residing in Bergen Municipality |  | = 12), mean age (SD) = 13.90 (0.85), male = 20%, female = 80%<br>Online Psychoeducation Program + Exposure Program (online PE + EXP) (n=18), mean age (SD) = 14.23 (1.07), male = 15%, female = 85%<br>Waitlist (WL) (n = 11), mean age (SD) = 14.05 (0.89), male = 10%, female = 90% | criteria<br>Online PE + EXP): Participants completed an online psychoeducation program, educated about PSA and CBT using bitmoji avatars, along with the online exposure program<br>Waitlist (WL): Placed on a waitlist for three weeks before accessing psychoeducation program. |
| Minns et al., 2019 | RCT | Full-scale study | Fear of spiders,<br>(Fear of Spiders Questionnaire (FSQ) | USA | 18-23 | 18-65 years<br>Assessed as unable to interact with a tarantula from the | 1 week | Immersive 3D Exposure-Based Treatment Group (n=38)<br>Psychoeducation -Only Control | Immersive 3D Exposure-Based Treatment Group: Six 5-minute 3D video exposures to spiders, utilizing the Oculus Rift DK1 virtual reality headset. |
|  |  |  |  |  |  | behavioral approach task (BAT)<br>Has not done exposure treatment for arachnophobia within three months. |  | Group (n=39) | Psychoeducation-Only Control Group:<br>Viewed a 30-minute documentary unrelated to spiders |
| Maskey et al., 2014 | Quasi-experimental (controlled trial) | Full-scale study | Specific phobias<br>(Spence Children's Anxiety Scale-parent version (SCAS-P) and child version (SCAS-C)) | UK | 7-13 years | Not mentioned | 2 weeks, 6 weeks, 6 months, 12-16 months | The study groups consist of individual participants with specific phobias or fears (n =9), mean age (SD) = 11.2 (2.0), boys = 100%, girls = 0%<br>No control group | Used a wrap-around virtual reality environment (VRE) called the "Blue Room," individualized exposure therapy |
| Nazligul et al., 2018 | RCT | Full-scale study | Public speaking anxiety<br>(Liebowitz | Turkey | 20-24 years | Score above 20 on LSAS subscales<br>Score above | Pre-Speech Anxiety<br>Peak | Experimental group (VREI group) (n=7)<br>Control group | VREI Group:<br>Used three-dimensional virtual environment, using real-life settings such as classrooms and |
|  |  |  | Social Anxiety Scale (LSAS) |  |  | threshold on the Social Interaction Anxiety Scale | Anxiety<br>Post-Speech Anxiety<br>Took 3 weeks | (CBT-based psychoeducation group) (n=7) | auditoriums, utilizing Oculus RIFT head mounted display<br>CBT-based Psychoeducation Group: Received CBT-based psychoeducation, involving traditional psychoeducational sessions discussing anxiety concepts and coping mechanisms |
| Rodero et al., 2022 | Quasi-experimental | Full-scale study | Public speaking anxiety<br>(Public Speaking Anxiety Scale (PSAS)) | Spain | 19-21 years | Not mentioned | Immediately after intervention | Experimental group (VRE) (n=50)<br>Control group (n=50) | Experimental Group: Trained with VR, including distractors (e.g., coughing, audience departure), conducting speech in front of virtual audience, utilizing the VR platform Psious<br>Control Group: Received traditional public speaking training without VR |
| Wechsler et al., 2024 | Quasi-experimental | Pilot study | Fear of spiders | Germany | 8-11 years | Ages 8–11 years | 1 week | All children went through VRET intervention | SpEYEders intervention: Gamified VR exposure used on a virtual spider, |
|  |  |  | (Spider Phobia Questionnaire for Children and Adolescents (SPF-KJ)<br>(BAV-11) |  |  | with and without spider fear |  | (n=21)<br>No control group | triggering positive transformations.<br>Children rated reactions and completed homework involving real spider exposure, utilizing HTC VIVE Pro Eye head-mounted display |
| Whiteside et al., 2020 | Quasi-experimental | Feasibility study | Academic worry in the context of anxiety disorders<br>(subjective units of distress (SUDs)) | USA | 8-18 years | Anxiety diagnosis using the Mini International Neuropsychiatric Interview for Children and Adolescents (MINI-KID), must have begun parent-coached exposure therapy | Immediately after intervention | All participants went through VRET and verbal imaginary exposure (Verbal IE) (alternative therapy) (n=20), mean age (SD) = 13.80 (2.88), girls: 70%, no demographics on boys | VRET: Immersed participants in a simulated classroom scenario using a VR headset to provoke and reduce anxiety through repeated exposure<br>Utilized Google Pixel Android smartphone and Daydream Viewer<br>Verbal IE: writing and repeatedly reading a personalized anxiety-inducing narrative |
| Porras-Garcia et | RCT | Feasibility preliminary | Eating disorder, | Spain | 14-18 years | 14-18 years | 5 weeks | VRET group (no | VRET group: Participants interacted |
| al., 2020 |  | y findings study | anorexia nervosa<br>(EDI-3: EDI-DT (drive for thinness), EDI-BD (body dissatisfaction)) |  |  | Patients from Eating Disorders Unit of the Hospital Sant Joan de Deu<br>Diagnosis of anorexia nervosa |  | demographics mentioned) (n=9)<br>Control group (no demographics mentioned) (n=8) | with virtual avatar, avatar's BMI increased, participants asked to focus on certain parts of avatar's body<br>VR head mounted display (HTC-VIVE) utilized<br>Control group: received CBT |
| Sulter et al., 2021 | Quasi-experimental | Proof of concept study | Public speaking anxiety<br>(Personal Report of Public Speaking Apprehension questionnaire) | Netherlands | 9-12 years | Children ages 9-12 years<br>Attending primary school | 1 week | SpeakApp-Kids! Condition (VRET), (n = 40), mean age (SD) = 10.72 (0.89), Boys: n= 23, Girls: n = 17<br>Control (n=49), mean age (SD) = 10.38 (0.84), Boys: n= 22, Girls: n= 27 | SpeakApp-Kids! Condition: children used the SpeakApp-Kids! virtual reality app, simulated a classroom environment with virtual audience.<br>utilized a Samsung Galaxy S7 Edge with a Samsung Virtual Reality Gear headset<br>Control: Practiced presentations at home, without virtual reality app |
| Ramsey et al., 2023 | Quasi-experimental | Feasibility study | Specific phobias (Clinical Global Impression □ Severity and Clinical Global Impression–Improvement (CGI □ S/CGI □ I) | USA | 9-13 years | Met diagnostic criteria for specific phobia | 1 week and 1 month | All participants exposed to VRET environment (n=3), mean age (SD) = 11.70 (2.09), females: n=2, males: n= 1<br>No control | Participants completed a single-session VR therapy with psychoeducation, and guided progression through challenging scenarios. Scenarios included a storm, dog, and spider environment for specific phobias<br>Utilized HTC Vive headset with headphones and the Virtually Better Inc. (VBI) exposure phobia suite |
| Tan et al., 2022 | Quasi-experimental | Feasibility study | Selective mutism<br>(selective mutism questionnaire ) | Singapore | 6-12 years | 6-12 years old, primary clinical diagnosis of selective mutism, completed at least 8 CBT session prior to study | Post VRET, 1 month, 3 months | All participants exposed to VRET environment (n=20), mean age (SD) = 8.45 (1.96), Boy: n = 11, Girls: n=9<br>No Control | Environment utilized school and classroom environments through interactive scenarios such as classroom participation and social interactions.<br>Custom virtual school environments displayed on TV screen |
| Farrell et al., 2021 | Quasi experimental, Multiple baseline case series | Pilot study, feasibility and preliminary efficacy study | Specific Phobia of dogs<br>(Spence Children's anxiety scale, child and parent version) | Australia | 8-12 years | 8 to 12 years with dog phobia, must have one parent willing to participate in treatment, stable medication dose | Post VRET, 1 month | All participants exposed to VRET environment (n=8), mean age (SD) = 10.25 (2.11), males: n=4, females: n=4<br>No control group | Participants were exposed to virtual scenarios with dogs, using hierarchy of exposure tasks while wearing a VR headset,<br>Utilized the Oculus Rift VR headset |
| Miller et al., 2023 | Quasi-experimental, single-arm open label study | Feasibility and preliminary efficacy | Depression (Center for Epidemiologic Studies Depression Scale, CES-D) | USA | 12-21 years | A PHQ-8 score of 10 or more, proficiency in English, access to mobile device. | Post intervention, 1 month | All participants exposed to VR-CBT intervention (n = 30) mean age (SD) = 17.03 (2.97), male: n=8, female: n = 21, non-binary: n = 1<br>No Control group | Used behavioral activation-based modules delivered via mobile app and immersive VR to teach coping strategies, enhance mood tracking, and support skill development<br>Utilized the Pico Interactive Goblin VR headset |
| De Luca et al., 2019 | Quasi-experimental, case study) | Feasibility study | Autism spectrum disorder (Gilliam Autism Rating Scale, | Italy | 16 years | Not mentioned | 2 months | Case study, participant went through CBT + VR training, (n=1), age = 16 years, male | Participant went through CBT and VR-CBT which included interactive tasks developed to enhance attention, spatial |
|  |  |  | GARS) |  |  |  |  |  | cognition, and adaptive behaviors through immersive, multisensory experiences<br>Utilized the BTS-Nirvana System, two-dimensional flat-screen projection system with optoelectronic infrared sensors |
| Kahlon et al., 2019 | quasi-experimental | Pilot study, feasibility study | Public speaking anxiety<br>(Public speaking anxiety scale (PSAS)) | Norway | 13-16 years | 13 to 16 years, fear of public speaking, score of 46-73 on PSAS, demonstrate functional impairment | Post intervention, 1 month, 3 months | All participants went under VRET (n=27), mean age (SD) = 14.22 (0.64), male: n=6, female: n = 21<br>No control group | VR environment simulating a classroom with virtual avatars, scenario included animated avatars seated at desks, an empty classroom, and a lobby, exposures had varying difficulty<br>Utilized Apple iPhone 7 paired with a premium Cardboard-style VR headset |
| Maskey et al., 2019 | RCT | Feasibility study | Specific Phobias (spence children's | UK | 7-14 years | 7-14 years, diagnosed with autism spectrum | 2 weeks, 6 months | Immediate treatment group (n=16), mean age (SD) = 10.84 | Immediate treatment group: received CBT combined with VR exposure therapy, |
|  |  |  | anxiety scale parent and child version, SCAS-P, SCAS-C) |  |  | disorder, can understand instructions, has specific phobias |  | (2.37), male: n= 13, female: n= 3<br>Control (delayed)group (n=16), mean age (SD) = 10.75 (1.79), male: n=12, female: n= 4 | Control (delayed) group: served as a no-treatment control group, completing assessments at 2 weeks and 6 months after randomization. After the 6-month assessment, received the same VR-based intervention as the immediate group<br>"Blue Room," VR utilized (no headset) |
| Porras-Garcia et al., 2020 | Quasi-experimental, case report | Feasibility study | Eating disorder, anorexia nervosa<br>(Eating disorder inventory (EDI3) -- ED-DT drive for thinness and ED-BD body dissatisfaction | Spain | 15 years | Not mentioned | Post-intervention, 5 months | Case study, participant went through VRET intervention (n=1), 15 years of age, female | Exposed to body mass index representation of their own virtual avatar<br>VR head mounted display (HTC-VIVE) utilized |
| Orena et al., 2023 | Quasi-experimental, case study | Feasibility study | Social anxiety related to Tourette syndrome plus (LSAS, Liebowitz Social Anxiety Scale) |  | 12 years | Not mentioned | Post-intervention, 1,3,6, and 10 months | Case study, participant went through both exposure therapy for social anxiety related to Tourette's syndrome, and VRET (n=1), 12 years, female | Comprehensive Behavioral Intervention for Tics (CBIT), addressing tics, with CBT targeting social anxiety, phobia, and obsessive-compulsive behaviors. VRET used for cognitive restructuring, assertive training, and mindfulness<br>Utilized the Oculus Quest 2, headset |

### 3.3 Mental disorder outcomes

#### 3.3.1 Anxiety disorders

A majority of mental disorders examined using VR-CBT were anxiety disorders (n=15) (Azimisefat et al., 2022; Farrell et al., 2021; Kahlon et al., 2019, 2023; Maskey et al., 2014, 2019; Minns et al., 2019; Nazligul et al., 2019; Orena & Baruffi, 2023; Ramsey et al., 2023; Rodero & Larrea, 2022; Sülter et al., 2022b; Y. L. Tan et al., 2025b; Wechsler et al., 2024; Whiteside et al., 2020). These studies included public speaking anxiety (n=5) (Kahlon et al., 2019, 2023; Nazligul et al., 2019; Rodero & Larrea, 2022; Sülter et al., 2022b) spider phobia (n=2) (Minns et al., 2019; Wechsler et al., 2024), dog phobia (n=1) (Farrell et al., 2021), acrophobia (n=1) (Azimisefat et al., 2022), social anxiety (n=1) (Kahlon et al., 2023) and selective mutism (n=1) (Y. L. Tan et al., 2025b). Notably, one study specifically examined academic worry as a dimension experienced by individuals with anxiety disorders (n=1) (Whiteside et al., 2020). Moreover, 3 studies measured several specific phobias within their participant samples, such as a ballon phobia, banana phobia, weather phobia, and various forms of animal phobias and social phobias (n=3) (Maskey et al., 2014, 2019; Ramsey et al., 2023). For these studies, each participant received customized VR scenarios suited to their specific phobia.

Validated scales and standardized instruments used to measure various anxiety disorder outcomes included the Severity Measure for Acrophobia (SMA), Visual Analogue Scale (VAS), Public Speaking Anxiety Scale (PSAS), Personal Report of Public Speaking Apprehension questionnaire, Fear of Spiders Questionnaire (FSQ), Spence Children’s Anxiety Scale (SCAS), Liebowitz Social Anxiety Scale (LSAS), Spider Phobia Questionnaire for Children and Adolescents (SPF-KJ), Clinical Global Impression-Severity and Clinical Global Impression–Improvement (CGI-S/CGI-I), subjective units of distress (SUDs), and the German BAV 3–11.

#### 3.3.2 Additional mental disorders

The remaining studies in this review focused on a range of mental disorders, such as depression (n=1)(Miller et al., 2023) (measured using the Center for Epidemiologic Studies Depression Scale, (CES-D)), autism spectrum disorder (ASD) (n=1) (L. Chu et al., 2023) (measured using the Gilliam Autism Rating Scale, GARS), ADHD (n=1) (L. Chu et al., 2023) (measured using the Attention-Deficit/Hyperactivity Disorder Rating Scale-IV (ADHD-RS-IV)), and Anorexia Nervosa (n=2) (Porras-Garcia, Serrano-Troncoso, Carulla-Roig, Soto-Usera, Ferrer-Garcia, Fernandez-Delcastillo Olivares, et al., 2020; Porras-Garcia, Serrano-Troncoso, Carulla-Roig, Soto-Usera, Ferrer-Garcia, Figueras-Puigderrajols, et al., 2020) (measured using the Eating disorder inventory (EDI-3)).

### 3.4 VR-CBT interventions

Within the broader category of VR-CBT interventions, 17 studies specifically employed VRET, a subset focused on exposure-based techniques (Azimisefat et al., 2022; L. Chu et al., 2023; De Luca et al., 2021; Farrell et al., 2021; Kahlon et al., 2019, 2023; Maskey et al., 2014, 2019; Miller et al., 2023; Minns et al., 2019; Nazligul et al., 2019; Orena & Baruffi, 2023; Porras-Garcia, Serrano-Troncoso, Carulla-Roig, Soto-Usera, Ferrer-Garcia, Fernandez-Delcastillo Olivares, et al., 2020; Porras-Garcia, Serrano-Troncoso, Carulla-Roig, Soto-Usera, Ferrer-Garcia, Figueras-Puigderrajols, et al., 2020; Ramsey et al., 2023; Y. L. Tan et al., 2025b; Wechsler et al., 2024). VRET was utilized as treatment for specific phobias (n=2) (Farrell et al., 2021; Ramsey et al., 2023), acrophobia (n=1) (Azimisefat et al., 2022), spider phobia (n=2) (Minns et al., 2019; Wechsler et al., 2024), dog phobia (n=1) (Farrell et al., 2021), academic worry in the context of anxiety disorders (n=1) (Whiteside et al., 2020), selective mutism (n=1) (Y. L. Tan et al., 2025b), eating disorders (n=2) (Porras-Garcia, Serrano-Troncoso, Carulla-Roig, Soto-Usera, Ferrer-Garcia, Fernandez-Delcastillo Olivares, et al., 2020; Porras-Garcia, Serrano-Troncoso, Carulla-Roig, Soto-Usera, Ferrer-Garcia, Figueras-Puigderrajols, et al., 2020), social anxiety related to Tourette’s syndrome (n=1) (Orena & Baruffi, 2023), and public speaking anxiety (n=5) (Kahlon et al., 2019, 2023; Nazligul et al., 2019; Rodero & Larrea, 2022; Sülter et al., 2022b). The remaining 3 studies were classified generally as VR-CBT (L. Chu et al., 2023; De Luca et al., 2021; Miller et al., 2023), utilizing general CBT principles without a primary focus on exposure therapy. Studies utilizing VR-CBT included interventions designed for individuals with ASD (n=1) (De Luca et al., 2021), depression (n=1) (Miller et al., 2023), and ADHD (n=1) (C. Chu et al., 2021).

#### 3.4.1 VR environment construction

In VRET, participants were placed in a virtual environment tailored to their specific phobia or mental disorder condition, such as public speaking settings (n=5) (Kahlon et al., 2019, 2023; Nazligul et al., 2019; Rodero & Larrea, 2022; Sülter et al., 2022b), navigating heights (n=1) (Azimisefat et al., 2022) or interactions with feared animals or social scenarios (n=8) (Farrell et al., 2021; Maskey et al., 2014, 2019; Minns et al., 2019; Orena & Baruffi, 2023; Ramsey et al., 2023; Wechsler et al., 2024; Whiteside et al., 2020). Studies integrated multiple sessions of VRET, following an exposure hierarchy of increasing levels of exposure to stimuli. For instance, the study by Porras-Garcia et al., 2020 utilizing VRET for Anorexia Nervosa exposed participants to a progressively increasing body mass index of a virtual avatar tailored to resemble their own body to systematically target their symptoms such as a drive for thinness and body dissatisfaction (Porras-Garcia, Serrano-Troncoso, Carulla-Roig, Soto-Usera, Ferrer-Garcia, Fernandez-Delcastillo Olivares, et al., 2020). Studies examining public speaking anxiety incorporated variations in audiences to target anxiety symptoms, such as number of audience members (Nazligul et al., 2019; Rodero & Larrea, 2022), duration of speech given (Rodero & Larrea, 2022; Sülter et al., 2022b), audience mood (Rodero & Larrea, 2022; Sülter et al., 2022b), and distractors during presentations (Rodero & Larrea, 2022). In the study by Tan et al., 2022 examining selective mutism in a classroom setting, scenarios first involved minimal interactions with virtual players (i.e. Participant required to respond to attendance roll call) to more complex interactions (i.e. Participant prompted to ask a question to teacher and respond to certain requests) (Y. R. Tan et al., 2022).

Moreover, 13 studies utilizing VRET interventions integrated performance feedback after completing varying levels to difficulty before moving onto the next exposure level (L. Chu et al., 2023; Farrell et al., 2021; Kahlon et al., 2019, 2023; Maskey et al., 2019; Minns et al., 2019; Nazligul et al., 2019; Orena & Baruffi, 2023; Porras-Garcia, Serrano-Troncoso, Carulla-Roig, Soto-Usera, Ferrer-Garcia, Figueras-Puigderrajols, et al., 2020; Ramsey et al., 2023; Y. L. Tan et al., 2025b; Wechsler et al., 2024; Whiteside et al., 2020). For example, Kahlon et al. (2023) employed a star rating system to determine the appropriate threshold level for tailoring the VRET intervention (Kahlon et al., 2023). For Maskey et al. (2019), participants progressed to a higher level of exposure after reporting consistently low anxiety levels. If anxiety increased from the previous level, therapists conducted relaxation techniques and anxiety monitoring (Maskey et al., 2019).

In studies using general VR-CBT, 2 studies reported participants engaged in interactive tasks in the virtual environment to improve cognitive function and teaching therapeutic strategies (C. Chu et al., 2021; De Luca et al., 2021). For example, Chu et al. (2023) utilized interactive game technology to increase children’s ability to process and respond to information (L. Chu et al., 2023). De Luca et al. (2019) interacted with the VR environment to perform motor and cognitive activities such as finding, counting, and moving objects (De Luca et al., 2021). The remaining study (n=1) utilizing VR-CBT had participants engaging in educational training (Miller et al., 2023). VR-CBT experiences consisted of training modules and educational videos about coping strategies, such as immersive videos on depression and behavioural activation, as well as mindful breathing exercises and meditations.

Additionally, 15% of VRET and VR-CBT interventions incorporated gamification by developing therapy as progressively challenging levels of exposure, including features such as virtual rewards or visually engaging graphics (L. Chu et al., 2023; Kahlon et al., 2023; Wechsler et al., 2024). For example, Wechsler et al. (2024) utilized gamified VRET for spider phobias by using animated spiders that danced to music and changed colors (Wechsler et al., 2024). Tan et al. (2022) reported that participants earned stars for progressing from nonverbal to verbal responses, which were tracked on a sticker chart and exchanged for small gifts after each VRET session (Y. R. Tan et al., 2022). Chu et al. (2023) examining ADHD utilized three interactive game scenarios where participants follow specific instructions to score points, with deductions for incorrect actions (L. Chu et al., 2023).

#### 3.4.2 VR technology utilization

A majority of studies employed wearable VR technologies (n=14) (Azimisefat et al., 2022; L. Chu et al., 2023; Farrell et al., 2021; Kahlon et al., 2019, 2019, 2023; Maskey et al., 2014; Minns et al., 2019; Nazligul et al., 2019; Porras-Garcia, Serrano-Troncoso, Carulla-Roig, Soto-Usera, Ferrer-Garcia, Fernandez-Delcastillo Olivares, et al., 2020; Porras-Garcia, Serrano-Troncoso, Carulla-Roig, Soto-Usera, Ferrer-Garcia, Figueras-Puigderrajols, et al., 2020; Ramsey et al., 2023; Y. L. Tan et al., 2025b; Wechsler et al., 2024; Whiteside et al., 2020), including VR glasses (n=1) (L. Chu et al., 2023), and VR headsets/headmounted displays (n=13) (Azimisefat et al., 2022; Farrell et al., 2021; Kahlon et al., 2023, 2023; Maskey et al., 2014; Minns et al., 2019; Nazligul et al., 2019; Porras-Garcia, Serrano-Troncoso, Carulla-Roig, Soto-Usera, Ferrer-Garcia, Fernandez-Delcastillo Olivares, et al., 2020; Porras-Garcia, Serrano-Troncoso, Carulla-Roig, Soto-Usera, Ferrer-Garcia, Figueras-Puigderrajols, et al., 2020; Ramsey et al., 2023; Y. L. Tan et al., 2025b; Wechsler et al., 2024; Whiteside et al., 2020). A total of n = 4 studies using VR headsets incorporated smartphones into their models such as Samsung, Google, and Apple products to display VR environments (Kahlon et al., 2019; Miller et al., 2023; Minns et al., 2019; Whiteside et al., 2020). The remaining studies utilized non-wearable technology, such as computer desktop projections (n=3) (C. Chu et al., 2021; Maskey et al., 2014, 2019), and TV projections (n=2) (C. Chu et al., 2021; Minns et al., 2019). One study utilized a VR phobia suite (n=1) (Ramsey et al., 2023), described as a multi-sensory VR exposure technology.

Most VR technology brands included Oculus technology (n = 6) (Farrell et al., 2021; Kahlon et al., 2019, 2023; Minns et al., 2019; Nazligul et al., 2019; Ramsey et al., 2023) and HTC Vive (n = 4) (Maskey et al., 2019; Porras-Garcia, Serrano-Troncoso, Carulla-Roig, Soto-Usera, Ferrer-Garcia, Fernandez-Delcastillo Olivares, et al., 2020; Rodero & Larrea, 2022; Wechsler et al., 2024) technologies as the primary platforms for delivering VR-CBT and VRET interventions. Other technology included various brands such as the Samsung gear VR (n=1) (Minns et al., 2019), Psious (n=1) (Rodero & Larrea, 2022), The Blue Room (n=2) (Maskey et al., 2014, 2019), Daydream viewer (n=1) (Whiteside et al., 2020), Pico Interactive Goblin VR (n=1) (Miller et al., 2023), and the BTS Nirvana System (n=1)(De Luca et al., 2021), Lenovo desktop screens (n=1) (L. Chu et al., 2023), and LCD and TV screens (n=2) (C. Chu et al., 2021; Minns et al., 2019).

**Table 4** presents the post-intervention means, standard deviations (SD), and effect sizes of the control and experimental groups in the meta-analysis. Azimisefat (2022) demonstrated a significant negative effect, with the experimental group scoring lower (M = 49.93, SD = 5.88) than the control group (M = 54.66, SD = 3.88). Similarly, Minns (2019) reported a significant negative effect (Hedge’s g = -0.818, 95% CI [-1.284, -0.353]), with the experimental group (M = 38.79, SD = 19.44) scoring lower than the control group (M = 56.92, SD = 24.12). In contrast, Kahlon (2023), Nazligul (2018), Rodero (2022), Sulter (2021), and Maskey (2019) reported non-significant findings. Rodero (2022) contributed the highest weight (24.950%) to the meta-analysis, reflecting a larger sample size and smaller variance.

**Table 4.**
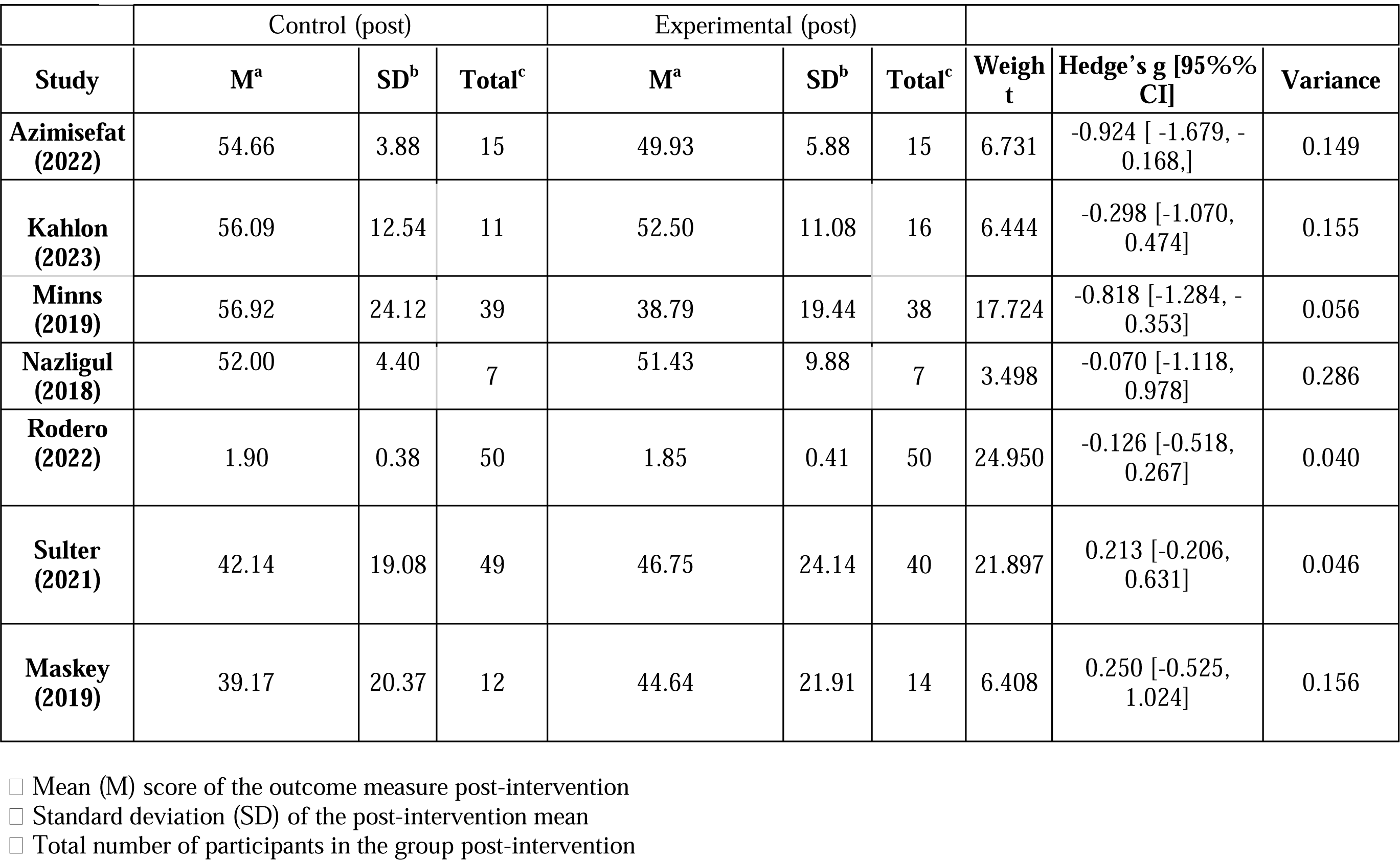
Post-intervention means, standard deviations (SD), and effect sizes of the control and experimental groups in the meta-analysis.

### 3.5 Efficacy Assessment through Meta-Analysis of Post-Intervention Means

**Figure 2** presents the forest plot of the effect sizes of five full-scale studies (Azimisefat et al., 2022; Kahlon et al., 2023; Minns et al., 2019; Nazligul et al., 2019; Rodero & Larrea, 2022) using Hedge’s g, and pooled estimate using a random effects model. The random-effects model yielded a pooled Hedge’s g of -0.46 (95% CI: [-0.84], [-0.09]), indicating a small to moderate negative effect size that is statistically significant. Moreover, **Figure 3** illustrates the forest plot of effect sizes for feasibility and full-scale studies using Hedge’s g, and pooled estimate using a random effects model. The meta-analysis pooled effect sizes from seven studies (Azimisefat et al., 2022; Kahlon et al., 2023; Maskey et al., 2019; Minns et al., 2019; Nazligul et al., 2019; Rodero & Larrea, 2022; Sülter et al., 2022b). However, the random effects model yielded a non-significant pooled effect size.

**Figure 1.**
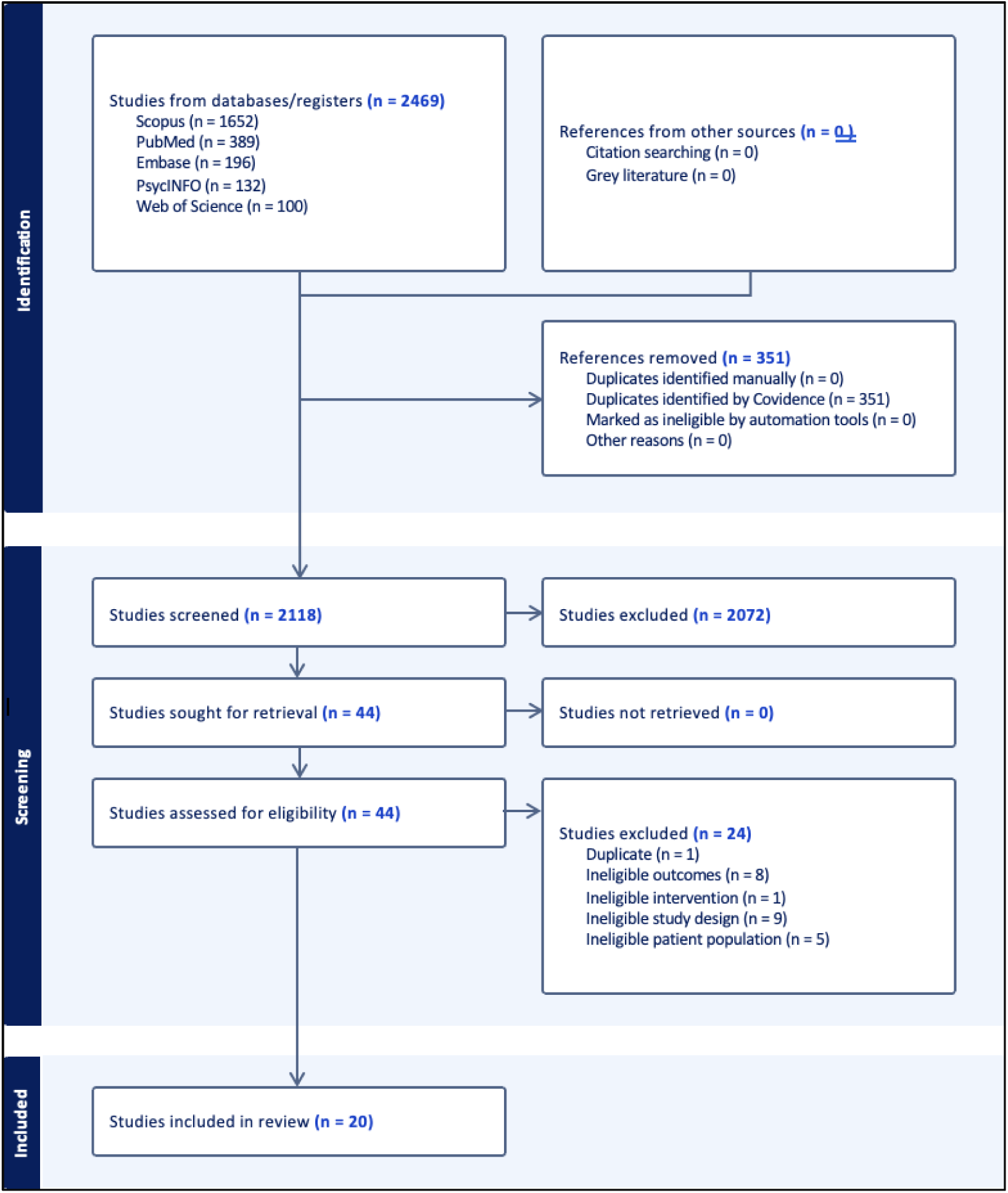
PRISMA flow diagram outlining the study selection process, including identification, screening, eligibility assessment, and inclusion of studies for the systematic review and meta-analysis.

**Figure 2.**
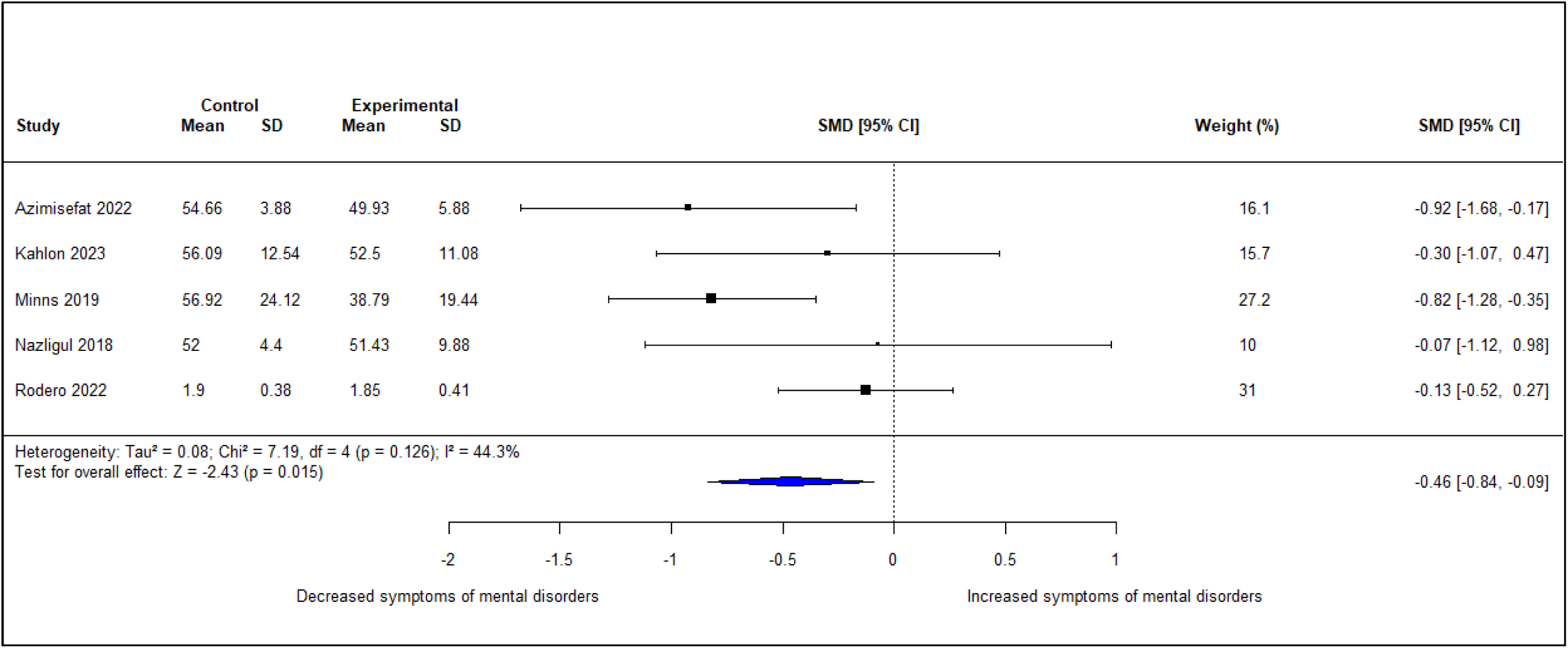
Forest plot of Hedge’s g effect sizes of full-scale studies and pooled estimate using a random-effects model.

**Figure 3.**
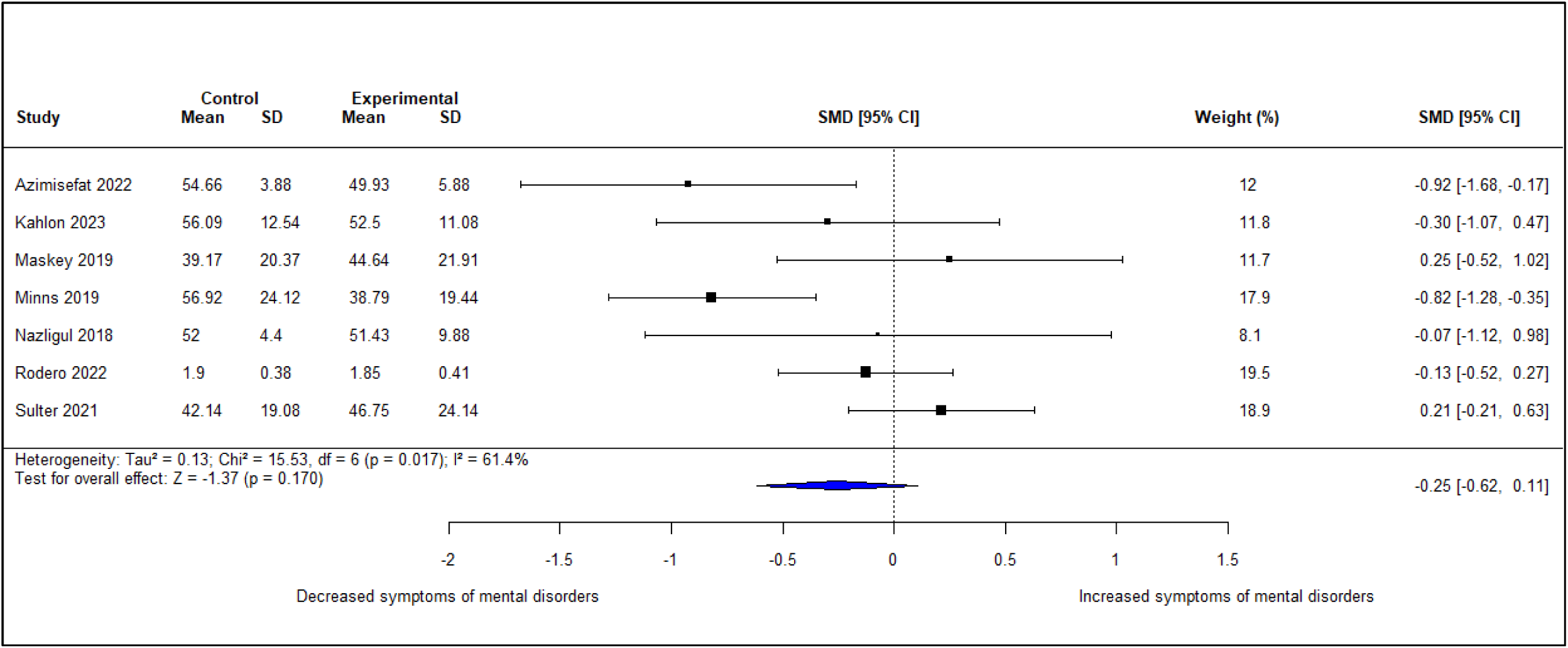
Forest plot of Hedge’s g effect sizes of feasibility and full-scale studies and pooled estimate using a random-effects model.

### 3.7 Risk of Bias Assessment

Risk of bias was assessed using both the JBI Critical Appraisal Tool for quasi-experimental studies **(Table 5)** and the Cochrane RoB 2 tool for RCTs **(Figure 4)**.

**Figure 4:**
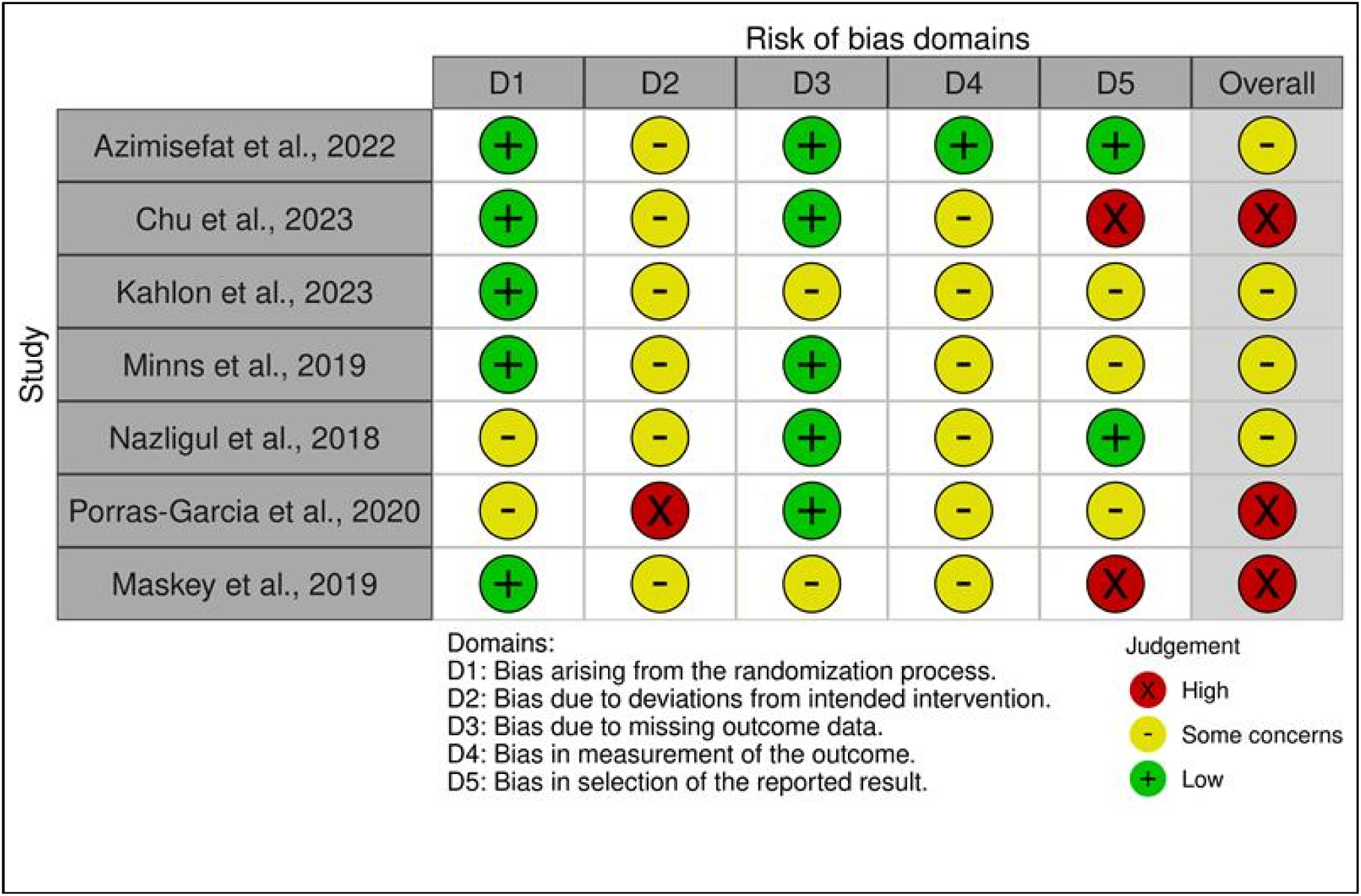
Cochrane RoB 2 Assessment.

**Table 5.**
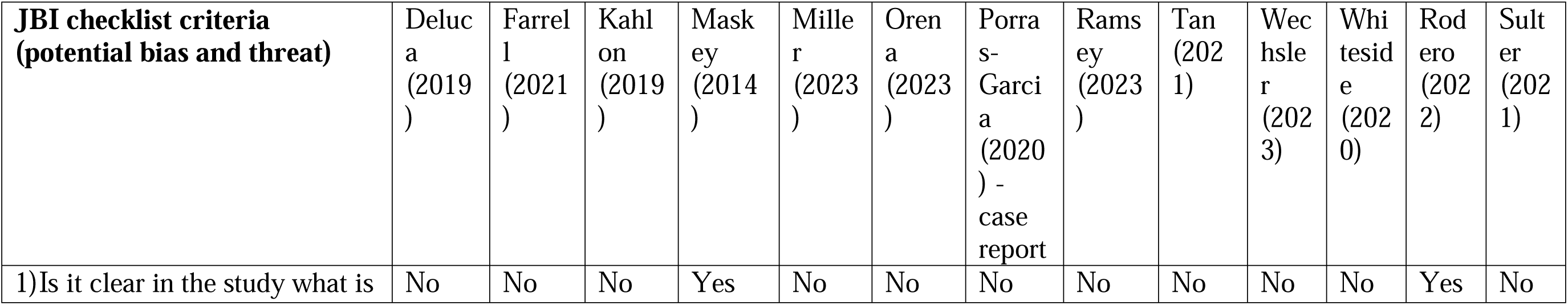
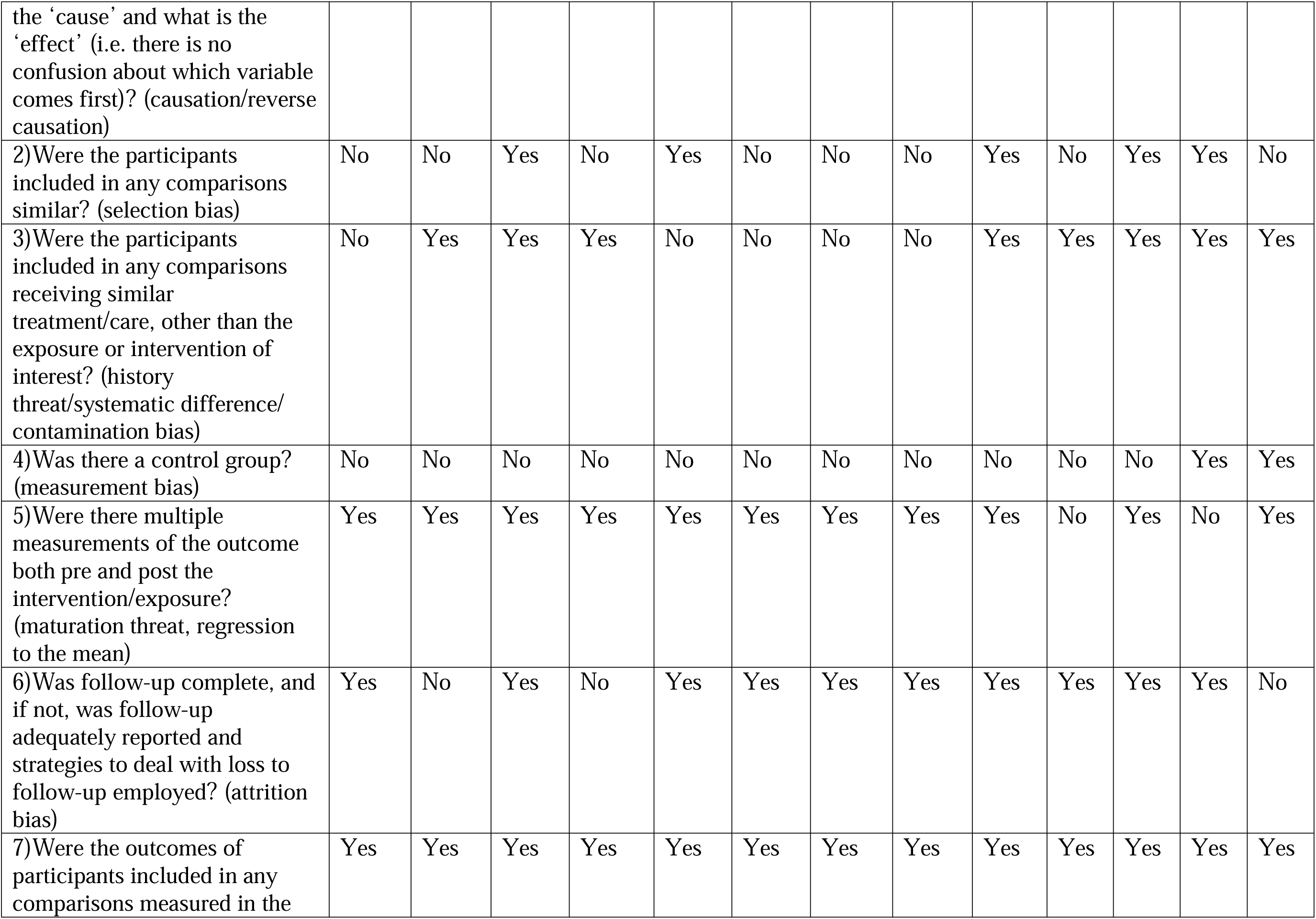
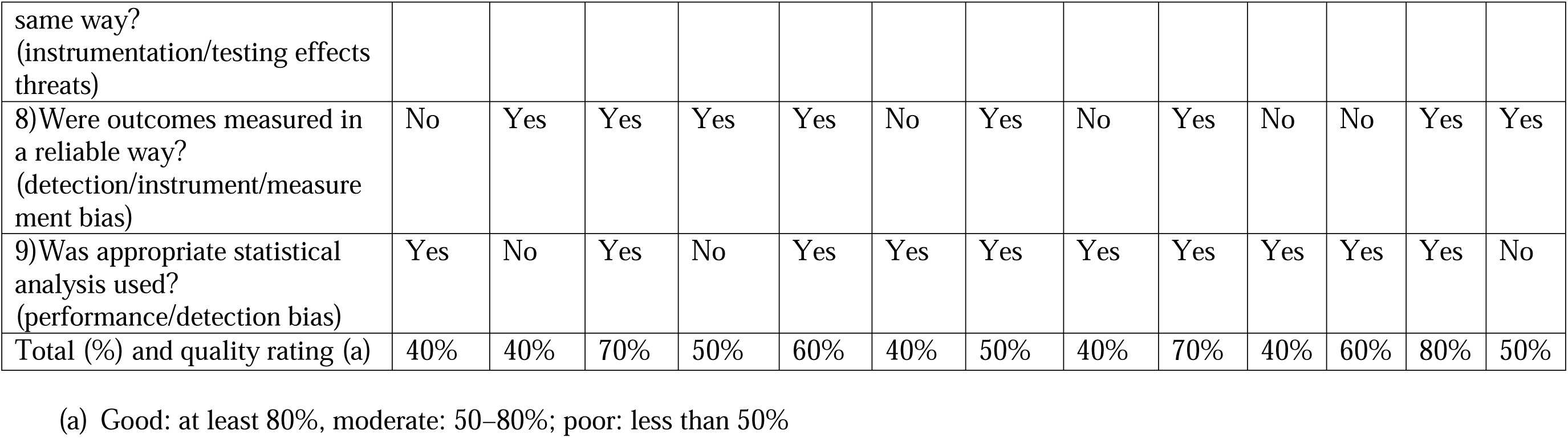
JBI critical appraisal tool for the assessment of risk of bias quasi-experimental studies.

Overall, the risk of bias across the included studies was moderate to high, with few studies demonstrating strong methodological quality and several studies exhibiting notable limitations in design, group comparability, and intervention fidelity. Out of the 20 studies assessed using either the JBI Critical Appraisal Tool (for 13 quasi-experimental studies) or the Cochrane RoB 2 tool (for 7 randomized studies), only one study (5%) met criteria for a “good” quality rating (Rodero & Larrea, 2022), while 11 studies (55%) were rated as moderate or had some concerns (Azimisefat et al., 2022; Kahlon et al., 2019, 2023; Maskey et al., 2014; Miller et al., 2023; Minns et al., 2019; Nazligul et al., 2019; Porras-Garcia, Serrano-Troncoso, Carulla-Roig, Soto-Usera, Ferrer-Garcia, Figueras-Puigderrajols, et al., 2020; Rodero & Larrea, 2022; Sülter et al., 2022b; Y. R. Tan et al., 2022; Whiteside et al., 2020), and eight studies (40%) were rated as poor or high risk (De Luca et al., 2021; Farrell et al., 2021; Orena & Baruffi, 2023; Ramsey et al., 2023; Wechsler et al., 2024). The JBI tool evaluated studies across nine domains, including clarity of cause-and-effect, group comparability, presence of a control group, similarity of care outside the intervention, outcome measurement, and appropriateness of statistical analysis. The Cochrane RoB 2 tool assessed bias across five domains: the randomization process (D1), deviations from intended interventions (D2), missing outcome data (D3), measurement of the outcome (D4), and selection of the reported result (D5).

Key methodological limitations were common across both types of studies. Among quasi-experimental studies, 84.6% lacked a clearly established cause-and-effect relationship and a control group (De Luca et al., 2021; Farrell et al., 2021; Kahlon et al., 2019; Miller et al., 2023; Orena & Baruffi, 2023; Porras-Garcia, Serrano-Troncoso, Carulla-Roig, Soto-Usera, Ferrer-Garcia, Figueras-Puigderrajols, et al., 2020; Ramsey et al., 2023; Y. R. Tan et al., 2022; Wechsler et al., 2024; Whiteside et al., 2020), and 61.5% of the studies involved non-comparable participant groups (De Luca et al., 2021; Farrell et al., 2021; Maskey et al., 2014; Orena & Baruffi, 2023; Porras-Garcia, Serrano-Troncoso, Carulla-Roig, Soto-Usera, Ferrer-Garcia, Figueras-Puigderrajols, et al., 2020; Ramsey et al., 2023; Sülter et al., 2022b; Wechsler et al., 2024). Additionally, 38.5% of the quasi-experimental studies did not ensure participants received similar care beyond the intervention (De Luca et al., 2021; Miller et al., 2023; Orena & Baruffi, 2023; Porras-Garcia, Serrano-Troncoso, Carulla-Roig, Soto-Usera, Ferrer-Garcia, Figueras-Puigderrajols, et al., 2020; Ramsey et al., 2023). Among randomized studies, 42.9% were rated as having a high overall risk of bias—primarily due to deviations from intended interventions (D2) and selective reporting (D5) (L. Chu et al., 2023; Maskey et al., 2019; Porras-Garcia, Serrano-Troncoso, Carulla-Roig, Soto-Usera, Ferrer-Garcia, Fernandez-Delcastillo Olivares, et al., 2020). The remaining 57.1% were judged to have “some concerns” (Azimisefat et al., 2022; Kahlon et al., 2023; Minns et al., 2019; Nazligul et al., 2019), particularly in bias due to deviations from intended interventions (D2) and bias in measurement of outcomes (D4), such as insufficient blinding of outcome assessors or reliance on self-reported measures without validation. Fewer issues were observed in randomization (D1) and missing outcome data (D3). Specifically, most randomized studies demonstrated adequate random sequence generation and allocation concealment, minimizing the risk of selection bias in D1. Similarly, for D3, the majority of studies had complete outcome data or used appropriate methods to handle missing data.

Despite these limitations, several methodological strengths were identified. Among quasi-experimental studies, outcome measurement was consistent in 100% of cases, with 84.6% of the studies using both pre- and post-intervention measurements (De Luca et al., 2021; Farrell et al., 2021; Kahlon et al., 2019; Maskey et al., 2014; Miller et al., 2023; Orena & Baruffi, 2023; Porras-Garcia, Serrano-Troncoso, Carulla-Roig, Soto-Usera, Ferrer-Garcia, Figueras-Puigderrajols, et al., 2020; Ramsey et al., 2023; Sülter et al., 2022b; Y. R. Tan et al., 2022; Whiteside et al., 2020)and 61.5% demonstrating reliable outcome measurement (Farrell et al., 2021; Kahlon et al., 2019; Maskey et al., 2014; Miller et al., 2023; Porras-Garcia, Serrano-Troncoso, Carulla-Roig, Soto-Usera, Ferrer-Garcia, Figueras-Puigderrajols, et al., 2020; Rodero & Larrea, 2022; Sülter et al., 2022b; Y. R. Tan et al., 2022). Additionally, 76.9% of the quasi-experimental studies reported adequate follow-up and used appropriate statistical analyses (De Luca et al., 2021; Kahlon et al., 2019; Miller et al., 2023; Orena & Baruffi, 2023; Porras-Garcia, Serrano-Troncoso, Carulla-Roig, Soto-Usera, Ferrer-Garcia, Figueras-Puigderrajols, et al., 2020; Rodero & Larrea, 2022; Sülter et al., 2022b; Y. R. Tan et al., 2022). For RCTs, key strengths included robust randomization procedures (D1) in 85.7% of studies (Azimisefat et al., 2022; L. Chu et al., 2023; Kahlon et al., 2023; Maskey et al., 2019; Minns et al., 2019) and low rates of missing outcome data (D3) in 71.4% of cases (Azimisefat et al., 2022; L. Chu et al., 2023; Minns et al., 2019; Nazligul et al., 2019; Porras-Garcia, Serrano-Troncoso, Carulla-Roig, Soto-Usera, Ferrer-Garcia, Fernandez-Delcastillo Olivares, et al., 2020). These features enhanced internal validity by reducing selection and attrition biases.

Taken together, the combined findings from both assessment tools indicate that while both study types demonstrated areas of methodological rigor, significant limitations across key domains of bias, especially intervention fidelity and selective reporting reduce the overall reliability and strength of the evidence base.

## 4.0 Discussion

This systematic review and meta-analysis examined the impact of VR-CBT on mental disorders among children and youth. Our main findings align with our initial hypothesis, that VR-CBT shows a significantly greater effect in reducing symptoms associated with mental disorders in children and youth compared to traditional therapies and controls (e.g., no treatment) at post-intervention. However, these findings were only applicable when analyzing full-scale studies. Although our inclusion criteria encompassed all mental disorders, only studies targeting anxiety-related conditions (specific phobias and public speaking anxiety) provided sufficient data for quantitative synthesis in our meta-analysis. These findings are consistent with previous systematic reviews and meta-analyses examining the effects of VR-based interventions on various mental disorders among children and youth, showing VR interventions to be effective for pain and anxiety, anxiety disorders, psychological distress (Kelson et al., 2021) and ADHD (Romero-Ayuso et al., 2021). For procedural anxiety among children and youth, results from a meta-analysis show VRET to be effective in lowering anxiety during medical procedures (Hu et al., 2024). Similar benefits have been observed in adult populations, where VR interventions have demonstrated effectiveness in addressing conditions such as anxiety disorders (Rejbrand et al., 2023), various phobias (Freitas et al., 2021), and PTSD (Gonçalves et al., 2012). For instance, a meta-analysis by Tan et al., 2024 shows VRET demonstrates greater effectiveness than waitlist controls in reducing social anxiety symptoms among adults (Y. L. Tan et al., 2025a).

The greater reduction in mental disorder symptoms in VR-CBT full-scale interventions can be explained by its ability to enable more timely treatment for mental health disorders compared to traditional CBT (Wu et al., 2021). Unlike traditional CBT, which often depends on "homework" assignments requiring patients to independently engage with fearful situations or apply therapeutic strategies in real-world situations (Jungbluth & Shirk, 2013; Kazantzis et al., 2010), VR-CBT integrates core components of CBT directly into the therapeutic sessions, allowing for real-time practice and application of therapeutic techniques. This was a consistent mechanism across all five full-scale studies in our meta-analysis: VR-CBT enabled real-time engagement with feared stimuli during therapy sessions. For instance, Rodero et al. (2022) and Nazligul et al. (2018) implemented VR-CBT public speaking interventions, allowing participants to deliver speeches within virtual environments featuring distractors, such as audience and mobile phone disruptions (Nazligul et al., 2019; Rodero & Larrea, 2022). These features allowed participants to engage with socially evaluative stimuli in-session, under controlled conditions (Nazligul et al., 2019; Rodero & Larrea, 2022). For Kahlon et al. (2023), adolescents participated in gamified VR exposure therapy enabling speech practice in front of a simulated audience, resulting in significant reductions in public speaking anxiety (Kahlon et al., 2023). Similarly, Azimisefat et al. (2022) exposed adolescent girls with acrophobia to height-related virtual scenarios (i.e., balconies and staircases) within a structured treatment framework (Azimisefat et al., 2022). Minns et al. (2019) further demonstrated that a single session of immersive VR exposure targeting spider phobia yielded significant reductions in fear (Minns et al., 2019). Collectively, these findings suggest that the therapeutic efficacy of VR-CBT may be driven by its capacity to enable real-time, therapist- or program-guided confrontation with anxiety-provoking stimuli, as opposed to delayed traditional CBT homework assignments. This approach can allow structured and consistent practice under the supervision of trained professionals, enhancing the effectiveness of, and adherence to the interventions.

Moreover, VR-CBT interventions can help mitigate the stigma associated with seeking mental health support (Seuling et al., 2024; Wu et al., 2021), which may improve accessibility for patients who might otherwise avoid treatment due to fear of judgment. Conducted within a confidential virtual environment, VR-CBT allows patients to engage in therapeutic exercises without the risk of potentially embarrassing or uncomfortable real-world exposures, providing a safe space to address their mental disorders (Fink et al., 2009; Schneier & Goldmark, 2015). For instance, Kahlon et al. (2023) provided patients with a gamified, self-guided VR program where they could repeatedly perform public-speaking in front of a virtual audience without the risk of social judgment from an in-person live audience (Kahlon et al., 2023). Similarly, Rodero et al. (2022) and Nazligul et al. (2018) created immersive VR environments with realistic social cues, such as audience noise or distractions to elicited anxiety, but within virtual simulations that did not involve real individuals (Nazligul et al., 2019; Rodero & Larrea, 2022). When using VR-CBT for specific phobias, Azimisefat et al. (2022) and Minns et al. (2019) allowed participants to engage in height- and spider-related exposures without direct contact with feared stimuli (Azimisefat et al., 2022; Minns et al., 2019). These applications underscore VR-CBT’s potential as a practical entry point into treatment for individuals with high avoidance, enabling earlier therapeutic engagement than conducting in vivo exposure alone.

Given the wide age range of children and youth included in our systematic review and meta-analysis, VR-CBT is shown to offer therapeutic engagement across age cohorts. For younger child cohorts of approximately ages 3-12 years, VR can be particularly engaging by leveraging their natural affinity for imaginative play and creativity. Children thrive in active, hands-on experiences that stimulate their curiosity and encourage exploration (Sobel & Letourneau, 2018). For instance, Wechsler et al. (2024) showed that children with a spider phobia responded well to a gamified VR-CBT intervention using playful mechanisms, including animating spiders to dance, diminish in size, or transform into different colors, to induce positive affect during spider exposure (Wechsler et al., 2024). Moreover, Chu et al. (2023) showed children engaged in game-like tasks, such as stepping on animated balloons or fish, that encouraged movement, exploration, and sensory-motor interaction. This format fostered strong engagement and compliance, leading to significant improvements in sensory processing, social relating, and response inhibition by leveraging children’s natural inclination toward play (L. Chu et al., 2023). Moreover, previous research suggests that engaging with immersive and stimulating environments can replenish cognitive resources and sustain focus (Yun et al., 2020). A scoping review by Cushnan et al., 2024 highlights integrating immersive tools in clinical mental health practices has shown promise in making therapeutic interventions more engaging, thereby improving adherence and outcomes (Cushnan et al., 2024). By providing a visually dynamic and interactive environment, VR-CBT can enhance motivation and adherence compared to verbally intensive CBT approaches that younger populations may find difficult.

Furthermore, VR-CBT can also be engaging for older youth cohorts (i.e., ages 16-24), who have grown up with technological advancement and heavily integrate technology as a crucial part of their daily lives (Haddock et al., 2022). Studies show that youth are highly receptive to technology-based interventions (Chen et al., 2024), often favoring them for their engaging format, flexibility, and convenience. Similarly, our systematic review and meta-analysis included studies that relied on immersive tools including the Oculus or HTC Vive headsets, smartphone-based VR platforms, or desktop projections. Within the broader scope of digital health, youth also show to prefer mobile health (mHealth) interventions over traditional in-person healthcare approaches (Radovic et al., 2018; Shapiro et al., 2008; Woolford et al., 2010). Moreover, a meta-analysis by Chen et al., 2024 showed digital technology interventions play an important role in addressing mental health challenges (Chen et al., 2024). In contrast to traditional therapies which rely on in-person sessions and may feel disconnected from their digital routines (Berardi et al., 2024; Harty et al., 2023), VR-CBT leverages their comfort and fluency with digital technology, potentially making it a more engaging and accessible therapeutic option.

In contrast to the reduction in mental disorder symptoms utilizing VR-CBT among full-scale studies, the inclusion of feasibility studies in our analysis resulted in a non-significant outcome. This difference in outcomes may be due to the unique characteristics of feasibility studies, which primarily evaluate the feasibility and implementation of study interventions as opposed to providing definitive conclusions on an intervention’s efficacy (Orsmond & Cohn, 2015). Moreover, feasibility studies often employ less rigorous controls, such as variations in randomization, comparator group design, and blinding (Orsmond & Cohn, 2015). These differences in methodology compared to full-scale studies could have affected the overall effect size, contributing to the non-significant outcome that was observed. Conducting more full-scale studies is essential to strengthening the literature on VR-CBT among children and youth, generating greater insights into its efficacy.

The current evidence on VR-CBT appears promising in addressing not only the overall effectiveness of the approach in comparison with traditional therapies, but also in bridging the critical access gaps among children and youth in obtaining timely mental health care (Azimisefat et al., 2022; Kahlon et al., 2023; Minns et al., 2019; Nazligul et al., 2019; Rodero & Larrea, 2022). However, some key recommendations can enhance both the effectiveness of VR-CBT interventions as well as the access to care for mental disorders among children and youth:

**1) mHealth platforms:** Integrating VR-CBT into mHealth platforms has the potential for on demand access to VR-CBT, promoting regular engagement outside of structured clinical settings. In a study conducted by Wiederhold et al., 2014, clinically tested VR environments were adapted for use on a mHealth phone application for chronic pain, resulting in patients reporting reduced pain and stress and an increase in relaxation (Wiederhold et al., 2014). Moreover, a systematic review examined mHealth and VR-based interventions for individuals with Alzheimer’s disease and found that these tools led to improvements in memory, cognition, mental health, and overall quality of life. The efficacy of this VR intervention can be attributed to mHealth’s ease of use, ability to use in daily routines, and scalability without the need of complex hardware (Kruse et al., 2022). Such findings can suggest that similar adaptations can be transferable to mHealth for mental health interventions, which can increase the accessibility of VR-CBT interventions. Given the high digital engagement of youth populations (Radovic et al., 2018; Shapiro et al., 2008; Woolford et al., 2010), integrating VR-CBT with mHealth may be particularly well-suited to meeting their mental health needs in a format that is both accessible and familiar.
**2) Big data integration:** VR-based therapies can be combined with other complementary tools such as wearable devices (Stasolla, 2021). This multimodal integration could enhance therapeutic impact by providing complementary strategies, such as real-time physiological monitoring and personalized biofeedback to address mental disorders from multiple approaches (Stasolla, 2021). In aligning mental health therapy with the current digital transformation of health systems (Katapally, 2024) that takes into account citizen/patient generated big data to enable human-centred artificial intelligence solutions (Ibrahim et al., 2025; Patel et al., 2025), VR-CBT interventions can obtain patient big data from mobile health platforms to enable digital phenotyping for advanced prediction and prevention of mental health disorder symptoms (i.e., real-time data collection to adapt therapy so it is tailored to patient- specific needs). Digital phenotyping is the continuous, real-time collection and analysis of behavioral and physiological data from digital devices (Bufano et al., 2023; Oudin et al., 2023). In the context of mental disorders, digital phenotyping can allow for ongoing assessment of factors associated with mental disorder symptoms during treatment sessions, such as stress levels or heart rate (Bufano et al., 2023; Oudin et al., 2023). In addition to VR-CBT, utilizing digital phenotyping could allow for significant adjustments to therapeutic exposures based on a patients’ real-time data, thus improving personalization of this treatment. With health systems currently evolving toward data-driven, patient-centered models (Boer et al., 2023; Jilka & and Giacco, 2024; Zhang et al., 2025), integrating VR-CBT with digital phenotyping and mobile health data sources presents an opportunity to deliver more tailored and proactive mental health care.
**3) Digital citizen science**: Both retention and compliance are key challenges when using VR-CBT interventions as a treatment, particularly in RCTs. Yet, digital citizen science can offer a solution to these limitations, as they actively engage participants in the development of VR interventions (Katapally, 2019, 2020a; Katapally et al., 2018). Co-creation activities, such as developing VR scenarios or providing feedback on therapeutic features, can allow for stakeholders such as children, youth, and caregivers (i.e., parents or legal guardians) to feel a sense of empowerment and develop a greater sense of ownership in the intervention and research processes (Katapally, 2019, 2020a, 2020b). This participatory approach to research not only works to align with their preferences for interactive and personalized treatment but can also enhance their motivation and adherence as participants (Katapally, 2019, 2020a; Katapally et al., 2018; Katapally & Chu, 2020). Thus, future studies utilizing VR-CBT for mental disorders should explore the integration of digital citizen science methodologies which can enhance engagement, improve retention, and support the development of more user-responsive and sustainable VR-CBT interventions. This digital citizen science approach might be particularly useful in utilizing VR-based mental health solutions for prevention of mental health disorders among individuals who have not been diagnosed of serious mental health disorders (i.e., by engaging children early in their childhood to predict and prevent, and if necessary to manage and treat mental health disorders).

## 5.0 Strengths and Limitations

The systematic review and meta-analysis examined the impact of VR-CBT interventions for various mental disorders in children and youth. By including a diverse range of mental disorders, our analysis developed a thorough evaluation of VR-CBT’s effectiveness and its potential future applications in mental health care. To avoid unbiased results, this review incorporated all studies investigating VR-CBT in this population, regardless of their reported outcomes, reducing selective reporting.

However, an important limitation of this review is the moderate to high risk of bias identified across the included studies. For instance, only one study included in our findings met criteria for a good-quality rating, while a majority demonstrated methodological concerns, especially in with group comparability, presence of a control group, and intervention fidelity. Additionally, other issues included non-comparable groups in quasi-experimental studies and deviations from intended interventions in randomized trials. These limitations reduce the overall reliability and internal validity of the evidence and emphasize the need for future VR-CBT studies to employ more rigorous designs with improved bias control across key methodological domains.

Our systematic review and meta-analysis show that future research is needed to build on our findings in this study, by adding more full-scale studies with rigorous methodology. While our systematic review included 20 studies that met our inclusion criteria, our meta-analysis comprised of seven studies: five full-scale studies and two feasibility studies. This limitation in number of studies was due to the requirement for experimental and control group post-intervention means of validated scales and standardized instruments to be analysed, which were not available in a significant number of studies identified within the systematic review. Yet, this limitation highlights the emerging nature of this field, as professionals have a growing interest in utilizing VR-CBT as a mental disorder intervention for children and youth. New research can enhance our understanding into the contexts and mechanisms to which VR-CBT is most effective in reducing mental disorder symptoms among the child and youth populations. As VR-CBT continues to evolve, our foundational results can provide a framework for advancing future work involving VR-CBT.

Moreover, our systematic review and meta-analysis only included studies published in English, which may have excluded relevant studies written in other languages. Additionally, another limitation is the heterogeneity in participant characteristics, type of VR technology, follow-up durations, and mental disorders among the studies included. As well, the small number of studies included in our review presented challenges for conducting detailed subgroup analyses.

## 6.0 Conclusion

This systematic review and meta-analysis show the clinical potential of VR-CBT in addressing mental disorders among children and youth. Our findings show VR-CBT can significantly reduce symptoms of mental disorders in comparison with traditional therapies and/or control groups. VR-CBT can be used as an opportunity to change the current state of mental health care delivery for children and youth who are highly receptive to digital interventions. For instance, integrating VR-CBT into mHealth platforms and multimodal approaches could expand both the accessibility and impact of this intervention among youth. Future research should conduct future studies on VR-CBT with a greater sample size and involve youth in co-creation processes to ensure interventions are both effective and youth-centered. These advancements hold significant potential to transform mental health care to help address the growing mental health burden globally.

## Data Availability

All data produced in the present work are contained in the manuscript

## 7.0 Data Availability

Data used in this review, including standard deviations, means, 95% confidence intervals, and effect sizes, are publicly accessible. Data has been deposited in a publicly available repository to promote transparency and facilitate replicability of the findings. However, primary data was not collected as a part of this research.

## Notes

### Competing Interest Statement

The authors have declared no competing interest.

### Funding Statement

The study was funded by the Canada Research Chairs Program

